# Broad SARS-CoV-2 cell tropism and immunopathology in lung tissues from fatal COVID-19

**DOI:** 10.1101/2020.09.25.20195818

**Authors:** Suzane Ramos da Silva, Enguo Ju, Wen Meng, Alberto E. Paniz Mondolfi, Sanja Dacic, Anthony Green, Clare Bryce, Zachary Grimes, Mary Fowkes, Emilia M. Sordillo, Carlos Cordon-Cardo, Haitao Guo, Shou-Jiang Gao

**Affiliations:** Cancer Virology Program, UPMC Hillman Cancer Center and Department of Microbiology and Molecular Genetics, University of Pittsburgh School of Medicine, Pittsburgh; Department of Pathology, Molecular and Cell-Based Medicine, Icahn School of Medicine at Mount Sinai, New York; Department of Pathology, University of Pittsburgh School of Medicine, Pittsburgh; Tissue and Research Pathology Core, UPMC Hillman Cancer Center, University of Pittsburgh School of Medicine, Pittsburgh

**Keywords:** SARS-CoV-2, COVID-19, cell tropism, diffuse alveolar damage, thromboemboli, interleukin-6, inflammation, immunosuppression, immunofluorescence assay, immunohistochemistry

## Abstract

**Background:** Severe Acute Respiratory Syndrome Coronavirus-2 (SARS-CoV-2) infection in patients with Coronavirus Disease 2019 (COVID-19) prominently manifests with pulmonary symptoms histologically reflected by diffuse alveolar damage (DAD), excess inflammation, pneumocyte hyperplasia and proliferation, and formation of platelet aggregates or thromboemboli. However, the mechanisms mediating these processes remain unclear.

**Methods:** We performed multicolor staining for viral proteins, and lineage cell markers to identify SARS-CoV-2 tropism and to define the lung pathobiology in postmortem tissues from five patients with fatal SARS-CoV-2 infections.

**Findings:** The lung parenchyma showed severe DAD with thromboemboli in all cases. SARS-CoV-2 infection was found in an extensive range of cells including alveolar epithelial type II/pneumocyte type II (AT2) cells (HT2-280), ciliated cells (tyr-α-tubulin), goblet cells (MUC5AC), club-like cells (MUC5B) and endothelial cells (CD31 and CD34). Greater than 90% of infiltrating immune cells were positive for viral proteins including macrophages and monocytes (CD68 and CD163), neutrophils (ELA-2), natural killer (NK) cells (CD56), B-cells (CD19 and CD20), and T-cells (CD3ε). Most but not all infected cells were positive for the viral entry receptor angiotensin-converting enzyme-2 (ACE2). The numbers of infected and ACE2-positive cells correlated with the extent of tissue damage. The infected tissues exhibited low numbers of B-cells and abundant CD3ε^+^ T-cells consisting of mainly T helper cells (CD4), few cytotoxic T cells (CTL, CD8), and no T regulatory cell (FOXP3). Antigen presenting molecule HLA-DR of B and T cells was abundant in all cases. Robust interleukin-6 (IL-6) expression was present in most uninfected and infected cells, with higher expression levels observed in cases with more tissue damage.

**Interpretation:** In lung tissues from severely affected COVID-19 patients, there is evidence for broad SARS-CoV-2 cell tropisms, activation of immune cells, and clearance of immunosuppressive cells, which could contribute to severe tissue damage, thromboemboli, excess inflammation and compromised adaptive immune responses.

**Funding:** This work used the UPMC Hillman Cancer Center and Tissue and Research Pathology/Pitt Biospecimen Core shared resource, which is supported in part by award P30CA047904 from the National Cancer Institute, and by UPMC Hillman Cancer Center Startup Fund and Pittsburgh Foundation Endowed Chair in Drug Development for Immunotherapy to S.-J. Gao.

**HIGHLIGHTS:** We provide an atlas of lung immunopathology of fatal SARS-CoV-2 infections, revealing:

- Unexpected broad cell tropism and infection of parenchymal, endothelial and immune cells by SARS-CoV-2, which are associated with massive tissue damage and thromboemboli;
- Clearance of immunosuppressive T-regulatory cells, and suppression of B cells and cytotoxic T cells;
- Extensive infiltration and activation of immune cells;
- Pronounced IL-6 expression in all types of infected and uninfected cells.

**Research in context:** *Evidence before this study:* Pulmonary symptoms reflected by diffuse alveolar damage (DAD), excess inflammation, pneumocyte hyperplasia and proliferation, formation of platelet aggregates, and thromboemboli are the pathological features of COVID-19. However, the mechanisms mediating these processes have not been elucidated. We searched PubMed up to September 15, 2020 using the keywords “coronavirus disease 2019”, “COVID-19”, “SARS-CoV-2”, “cell tropism”, “cell markers”, “inflammation”, “interleukin 6”, “immune response”, “immune suppression”, “immunofluorescence” and “immunohistochemistry”, with no language restrictions. Single cell RNA sequencing (scRNA-seq) has revealed extensive expression of SARS-CoV-2 receptor angiotensin-converting enzyme-2 (ACE2) in a large variety of cell types. However, only low levels of SARS-CoV-2 infection have been detected in macrophages, neutrophils, type II pneumocytes (AT2), and goblet, club, ciliated and endothelial cells by scRNA-seq and immunohistochemistry. COVID-19 blood samples contain high levels of inflammatory cytokines including interleukin-6 (IL-6), high levels of monocytes and neutrophils, and depletion of lymphocytes. There is no information on the cell types infected by SARS-CoV-2 and extent of infection, the precise producing cells of inflammatory cytokines, and the status of immune cells in lungs from fatal COVID-19 patients.

*Added value of this study:* By multicolor staining for viral proteins and lineage markers in lung tissues from five fatal COVID-19 patients, we reveal SARS-CoV-2 infection in an extensive range of cells including type II pneumocytes (HT2-280), and ciliated (tyr-α-tubulin), goblet (MUC5AC), club-like (MUC5B) and endothelial cells (CD31 and CD34), which is correlated with the extent of DAD and thromboemboli. SARS-CoV-2 infection is found in greater than 90% of infiltrating immune cells, including macrophages and monocytes (CD68 and CD163), neutrophils (ELA-2), natural killer cells (CD56), B-cells (CD19 and CD20), and T-cells (CD3ε). Most but not all infected cells were positive for ACE2. There are abundant macrophages, monocytes, neutrophils and natural killer cells but low numbers of B-cells and abundant CD3ε^+^ T-cells consisting of mainly T helper cells (CD4), few cytotoxic T cells (CTL, CD8), and no T regulatory cell (FOXP3). Antigen presenting molecule HLA-DR of B and T cells was abundant in all cases. Robust IL-6 expression was present in most uninfected and infected cells, with higher expression levels observed in cases with more tissue damage.

*Implications of all the available evidence:* In lung tissues from severely affected COVID-19 patients, there is evidence for broad SARS-CoV-2 cell tropisms, hyperactive immune cells, and clearance of immune cells including immunosuppressive cells, which could contribute to severe tissue damage, thromboemboli, excess inflammation and compromised adaptive immune responses. These results have implications for development of treatments.

## Introduction

Coronavirus Disease 2019 (COVID-19) is a complex disease caused by Severe Acute Respiratory Syndrome Coronavirus-2 (SARS-CoV-2) infection.^1,2^ Multiple organs are affected, and severe lung damage is a prominent finding in fatal cases.^3-6^ Although dysregulated immune responses and excess inflammation are commonly observed in the lung tissues from these patients, the precise mechanism underlying the pulmonary pathology remains unclear.^4^

Single cell RNA sequencing (scRNA-seq) analysis of lung tissues from healthy subjects have revealed that many cell types express SARS-CoV-2 entry receptor and cofactors including angiotensin-converting enzyme-2 (ACE2), transmembrane serine protease 2 (TMPRSS2), and furin, that are involved in viral entry, suggesting susceptibility of these cells to infection.^7-10^ Furthermore, scRNA-seq analysis of bronchoalveolar lavage fluid (BALF), blood, oropharyngeal or lung tissues from COVID-19 patients has identified different types of SARS-CoV-2-infected cells, including macrophages, neutrophils, type II pneumocytes (AT2), and ciliated and endothelial cells.^11-14^ However, in general, these studies detected very low numbers of infected cells, which harbored low counts of viral genomes and transcripts.^11-16^ The reason for the discrepancy between the high numbers of cells expressing viral entry receptors/cofactors and the low numbers of infected cells detected even in COVID-19 patients with severe pulmonary disease remains unclear. It has been reported that the expression of ACE2, TMPRSS2 and furin is upregulated in macrophages, neutrophils, AT2 and ciliated cells in COVID-19 patients compared to healthy controls, and that type 1 interferons (IFNs) induce the expression of ACE2 in epithelial cells, hence increasing their susceptibility to infection.^17,18^ However, a recent study showed that type 1 IFNs only induced the expression of an ACE variant but not the ACE2 involved in viral entry.^19^ Furthermore, although immunohistochemistry (IHC) staining of lung tissues with antibodies detected SARS-CoV-2 spike (S1) protein or nucleocapsid (NC) protein in macrophages (cluster of differentiation 68^+^, CD68^+^ and CD183^+^), and AT2, ciliated, goblet, club and endothelial progenitor cells, the infected cells were often observed at low numbers, and the exact identity of many infected cells remain unknown.^20-24^ Questions remain regarding the SARS-CoV-2 targeted cell types, the percentages of the cells that are infected, and whether the extent of infection is correlated with the expression of viral entry factors and disease status.

Interleukin-6 (IL-6) is one of the most abundant cytokines detected in COVID-19 patients.^25^ The expression level of IL-6 has been correlated with patient prognosis.^26-28^ Treatment with IL-6 antagonists improved the survival and shortened the recovery time.^29-33^ However, the cell types responsible for increased IL-6 expression in the lung are poorly defined, and understanding the relationship among IL-6 expression, the extent of SARS-CoV-2 infection, and disease severity is incomplete.

In this study, we analyzed the expression of SARS-CoV-2 S1 and NC proteins in postmortem lung tissues from five severe COVID-19 patients with various degrees of lung damage. We performed multicolor immunofluorescence staining (IF) for the SARS-CoV-2 proteins, ACE2 protein as well as for lineage-restricted cell markers. We found broad and extensive SARS-CoV-2 infection in the lungs of these patients, and more infected cells were observed in more severe cases. Infected immune cell types were comprised of monocytes and macrophages (CD68^+^ or CD163^+^), neutrophils (ELA-2^+^), and natural killer (NK) (CD56^+^), B (CD20^+^), and T (CD3ε^+^, CD4^+^and CD8^+^) cells, including activated B and T (HLA-DR^+^) cells with many having near 100% of infection in severe cases. To our knowledge, this is the first direct visualization by IHC and IF of SARS-CoV-2 infection of neutrophils and different T cell subtypes. We simultaneously detected SARS-CoV-2 infection and ACE2 expression in AT2 pneumocytes, and club-like, goblet and endothelial cells. Finally, we found wide spread IL-6 expression in lung parenchyma involving most of the cells and cell types regardless of individual cell infection status.

## Methods

### COVID-19 lung tissue samples

Anonymized postmortem specimens were collected from five adults (4 male and 1 female) with fatal SARS-CoV-2 infection by the Autopsy Service of the Department of Pathology, Molecular and Cell-based Medicine at the Icahn School of Medicine at Mount Sinai. All 5 patients had been admitted because of symptomatic COVID-19 and a positive nasopharyngeal swab test for SARS-CoV-2 by real-time reverse-transcription polymerase-chain-reaction amplification (RT-PCR, cobas® 6800 system, RocheDiagnostics). Other clinical-pathologic findings are summarized in table S1. All autopsies were performed with written consent from the legal next-of-kin, and specimens were obtained per the Autopsy Service protocol.

Specimens obtained at autopsy do not meet the definition of a living individual per Federal Regulations 45 CFR 46.102, and as such, research using specimens obtained at autopsy does not meet the requirements for Institutional Review Board review or oversight under the Icahn School of Medicine Program for the Protection of Human Subjects. In addition, the Institutional Review Board of the University of Pittsburgh determined that the study is not research involving human subjects as defined by DHHS and FDA regulations and waived of ethical oversight (STUDY20050085).

### Hematoxylin-eosin (H&E) staining, immunohistochemistry (IHC), and immunofluorescence assay (IF)

Postmortem biopsies were fixed with 10% neutral buffered formalin and embedded in paraffin. Slides were stained with H&E for histological analyses. For IHC single staining (CD3, CD4, CD8, CD45, CD19, CD20 and FOXP3), the slides were deparaffinized at 60°C for 30 min and rehydrated using a standard histology protocol of 3 changes of xylene of 5 min each followed by 3 changes of ethanol 100%, 2 of ethanol 95% and 1 ethanol 70% for 1 min each, then rinsed in distilled water. Antigen retrieval was performed using citrate buffer (#S1699, Agilent Dako) in Decloaking chamber at 123°C for 2 min. The slides were stained using an Autostainer Plus (Agilent Dako) platform with TBS-T rinse buffer (#S3306, Agilent Dako). The IHC slides were treated with 3% hydrogen peroxide for 10 min. The primary antibodies were applied at room temperature for 30 min, followed by 30 min of secondary antibody Envision + Dual Link (# K4061, Agilent Dako) HRP polymer at room temperature. Slides were exposed to 3,3, Diaminobenzidine+ (# K3468, Agilent Dako) for 5 min, and counterstained with Hematoxylin (#K8018, Agilent Dako). For IF, slides were deparaffinized at 95°C for 10 min, followed by 3 washes of xylene for 5 min. Dehydration was performed with step-wise 10 min incubation of ethanol 100%, 95% and 75%, followed by water. Antigen retrieval used citrate buffer pH 6.0 on microwave for 3 min at maximum potency, followed by 15 min with 30% potency, and cooled down for 30 min at room temperature. Slides were treated for 1 h with 5% bovine serum albumin (BSA) solution. Primary antibodies were incubated overnight at 4°C, and secondary antibodies were incubated for 1 h at room temperature. Slides were treated with Vector TrueVIEW™ autofluorescence quenching (#SP-8400, Vector Laboratories) for 5 min followed by incubation with 4’,6-diamidino-2-phenylindole (DAPI) for 10 min. Table S2 summarizes all antibodies and dilutions used in the study.

## Results

All five cases showed various combinations of DAD, pulmonary thromboemboli and pulmonary consolidation (table S1, figure 1A and S1A). Case 4 had the most extensive and severe pathologic changes, including early exudative phase of DAD, vascular congestion and rare hyaline membranes. Air-spaces filled with blood were noted in cases 1, 2 and 4. The least dramatic changes were found in cases 2 and 3; both had incidental anthracosis. These findings are in agreement with previous descriptions of lung pathology in COVID-19 cases.^6,21,34^

**Figure 1:**
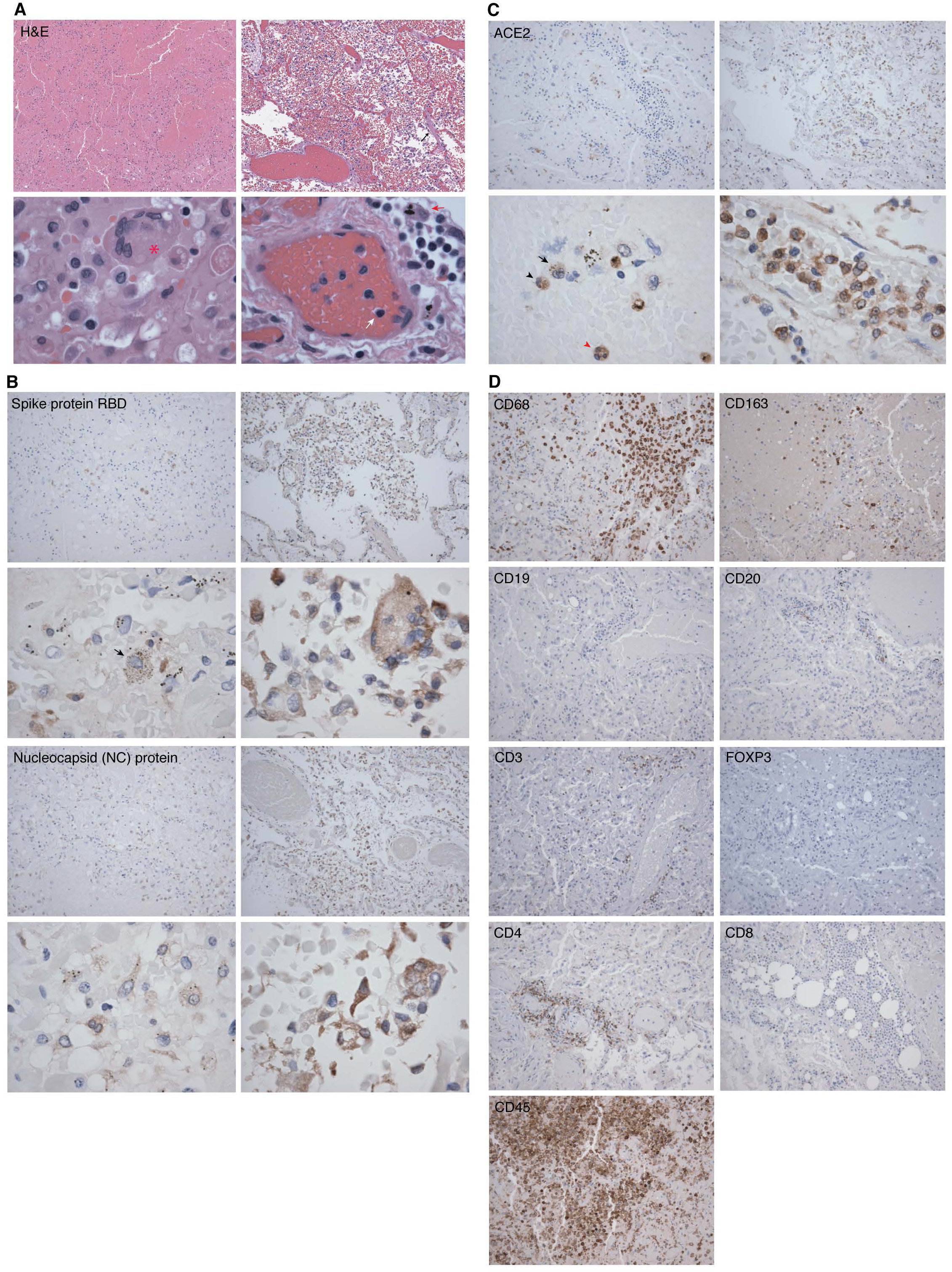
Representative hematoxylin-eosin (H&E) and immunohistochemistry (IHC) staining images of SARS-CoV-2 proteins and markers of immune cells in lung tissues from two COVID-19 patients. Shown are H&E images illustrating significant areas of lung tissues (Panel A, case 2 in left images and case 4 in right images). An image of case 2 showing lung parenchyma with hemorrhagic infarct in top image (100x). A Langhans giant cell is visible in bottom image (asterisk, 600x). An image of case 4 with an early exudative phase of DAD, vascular congestion and rare hyaline membranes in top image (black arrow, 100x), and infiltrations of lymphocytes (white arrow) and macrophages (red arrow) in bottom image (600x). Shown are IHC detection of SARS-CoV-2 infection using antibody against spike protein (receptor binding domain, RBD) and NC protein (100x) (Panel B, case 2 in left images and case 4 in right images). A macrophage infected by SARS-CoV-2 is visible in case 2 bottom image (black arrow, 600x). Case 2 has less positive cells compared to case 4 for both viral proteins. Shown are IHC detection of ACE2 protein expression in lung tissues (Panel C, case 2 in left images and case 4 in right images). Immune cells identified in case 2 in bottom image are a monocyte (black arrowhead), a macrophage (black arrow) and a neutrophil (red arrowhead), all expressing ACE2 protein (600x). Shown are IHC detection of markers of immune cells in a lung tissue from a COVID-19 patient (case 3) consisting of monocytes and macrophages (CD68^+^ and CD163^+^), B cells (CD19^+^ and CD20^+^), different markers of T cells including T cell receptor (CD3ε^+^), T regulatory cell (FOXP3), helper T cell (CD4^+^), cytotoxic T cell (CD8^+^), and lymphocyte common antigen (CD45^+^) (100x, Panel D).

Evidence of SARS-CoV-2 infection was detected by IHC with antibodies against the S1 protein receptor binding domain (RBD) and NC protein. All 5 cases were positive for SARS-CoV-2 proteins (figure 1B and S1B) with the widest distribution of infected cells observed in tissues from case 4 followed by cases 1 and 5. The fewest infected cells were observed in lung tissues from cases 2 and 3. Tissue damage was widespread in all cases. Case 5 had the most structurally preserved tissue specimen with infected cells present in patches, rather than throughout the lung specimen.

Consistent with the detection of viral proteins, in all cases the ACE2 protein, the main receptor for SARS-CoV-2^35,36^, was widely detected in different cell types including immune cells comprised of monocytes, macrophages, neutrophils and lymphocytes (figure 1C and S1C). The extent of ACE2 protein expression correlated with that of SARS-CoV-2 infection with cases 2 and 3 having the lowest numbers of cells expressing ACE2 protein (figure S1C).

Infiltration of immune cells is a common sequel to infection. We identified immune cells by staining for different cell markers by IHC. Cells positive for CD45 (leukocyte common antigen, LCA), a marker for most hematopoietic cells, was highly abundant in all cases (figure 1D and S2C). Abundant infiltrating monocytes and macrophages were detected in all cases using CD68 as a monocyte, pan-macrophage or M1 marker, and CD163 as a M2 cell marker. CD68^+^ cells were more abundant than CD163^+^ cells (figure 1D and S2A).

We identified B cells by staining for CD19 and CD20. Although there was a paucity of CD19^+^ cells, lung tissues from cases 1 and 2 showed pockets of infiltrating CD20^+^ cells, which appeared to be surrounding venous structures (figure 1D and S2B). Infiltration by T cell receptor (TCR) CD3ε^+^ cells, predominantly T CD4^+^ helper, and fewer T CD8^+^ cytotoxic cells, was detected in all cases (figure 1D and S2C). Interesting, all cases were negative for FOXP3, a marker for natural T regulatory (Treg) cells (figure 1D and S2C).

Since ACE2 protein expression was correlated with SARS-CoV-2 infection (figure 1B-C, and S1B-C), we performed dual IF staining for ACE2 and SARS-CoV-2 S1 protein (figure 2A and S3). Most infected cells expressed ACE2 protein but we also observed some ACE2-negative infected cells, which could be due to low expression level of ACE2 protein outside the detection range of the assay, downregulation of ACE2 protein expression at some stage(s) of SARS-CoV-2 infection, virus cell-to-cell spread, or presence of an alternative SARS-CoV-2 receptor. Consistent with our IHC findings, lung tissue specimens displaying a broader range of SARS-CoV-2-infected cell types, also had more ACE2-positive cells (figure S3). Cases 2 and 3 had the lowest numbers of ACE2-positive cells and the least infected cells.

**Figure 2:**
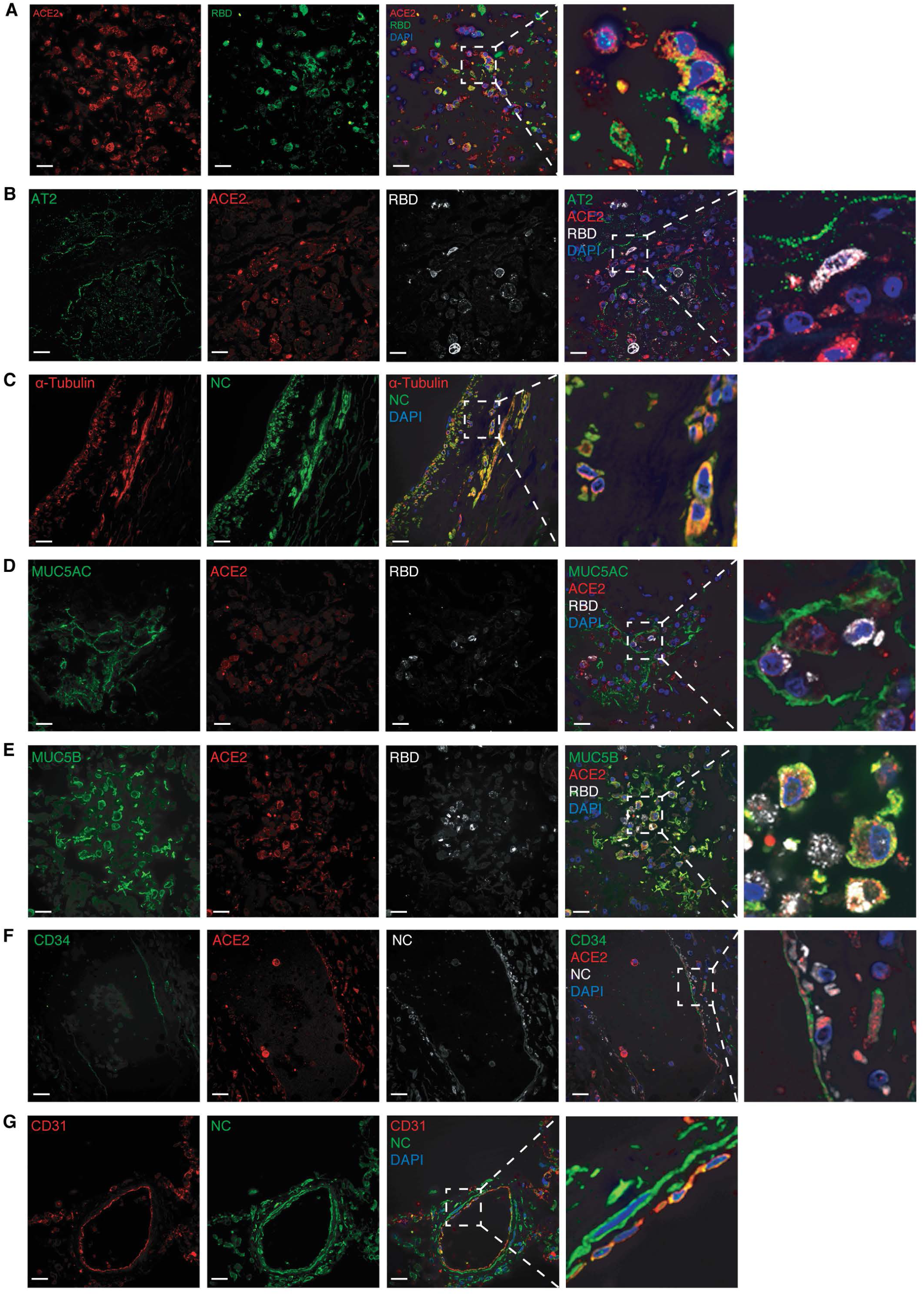
Representative images of multicolor immunofluorescence staining of ACE2, SARS-CoV-2 proteins and cellular markers in lung tissues from COVID-19 patients. Shown are ACE2 protein (pseudo color red) and SARS-CoV-2 S1 protein (RBD, pseudo color green) in case 4 (Panel A); Alveolar epithelial type II / pneumocytes type II cells (AT2) (pseudo color green), ACE2 (pseudo color red) and SARS-CoV-2 S1 protein (RBD, pseudo color white) in case 5 (Panel B); Tyr-α-tubulin (pseudo color red) and SARS-CoV-2 NC protein (pseudo color green) in case 5 (Panel C); MUC5AC (goblet cells, pseudo color green), ACE2 (pseudo color red) and SARS-CoV-2 S1 protein (RBD, pseudo color white) in case 4 (Panel D); MUC5B (club-like cells, pseudo color green), ACE2 (red) and SARS-CoV-2 S1 protein (RBD, pseudo color white) in case 4 (Panel E); CD34 (pseudo color green), ACE2 (red) and SARS-CoV-2 NC protein (pseudo color white) in case 5 (Panel F); and CD31 (pseudo color red) and SARS-CoV-2 NC protein (pseudo color green) in case 1. Nuclei were stained with DAPI (pseudo color blue) (Panel G).

Since we observed extensive damage to lung tissues (figure 1A and S1A), we performed triple-color staining for SARS-CoV-2 and ACE2 proteins in lung parenchymal cells including AT2 cells, ciliated cells (tyr-α-tubulin), goblet cells (MUC5AC), and club-like cells (MUC5B) in order to evaluate these cell types for viral infection (figure 2B-E and S4A-D). AT2 cells, which have been reported to be a major target of SARS-CoV-2 infection in lung tissues^17^, were shown both to express ACE2, and to be infected by SARS-CoV-2 (figure 2B and S4A). In sections of lung tissues from cases 2 and 3, AT2 cells were noted to have intact cell membranes, and less extensive infection by SARS-CoV-2. In contrast, in case 4, more extensive SARS-CoV-2 infection of AT2 cells was observed, along with ruptured cell membranes, possibly related to viral shedding (figure S4A).

Tyr-α-tubulin was used as a microtubule marker for identifying ciliated cells among others. SARS-CoV-2 extensively infected cells expressing tyr-α-tubulin, including ciliated cells identified by their morphology, in lung tissues from all cases (figure 2C and S4B).

The predominant mucins expressed in the lung are MUC5AC, mainly found in goblet cells, and MUC5B, mostly expressed in club-like cells. Both MUC5AC^+^ and MUC5B^+^ cells were infected by SARS-CoV-2 and expressed ACE2 (figure 2D-E and S4C-D). MUC5B^+^ cells were more abundant than MUC5AC^+^ cells. Previous studies have shown SARS-CoV-2 infection of small number of vascular endothelial cells in lung tissues from COVID-19 patients.^37^ We observed extensive SARS-CoV-2 infection and damage in CD34^+^ or CD31^+^ endothelial cells (figure 2F-G and S5A-B), which might be the cause of widespread microhemorrhages and infarction observed in these tissues.

Since we detected vast infiltrations by innate immune response cells, we examined SARS-CoV-2 infection in these cells. Monocytes and macrophages (CD68^+^ or CD163^+^) were widely infected by SARS-CoV-2 (figure 3A, B and E, and S6A-B).

**Figure 3:**
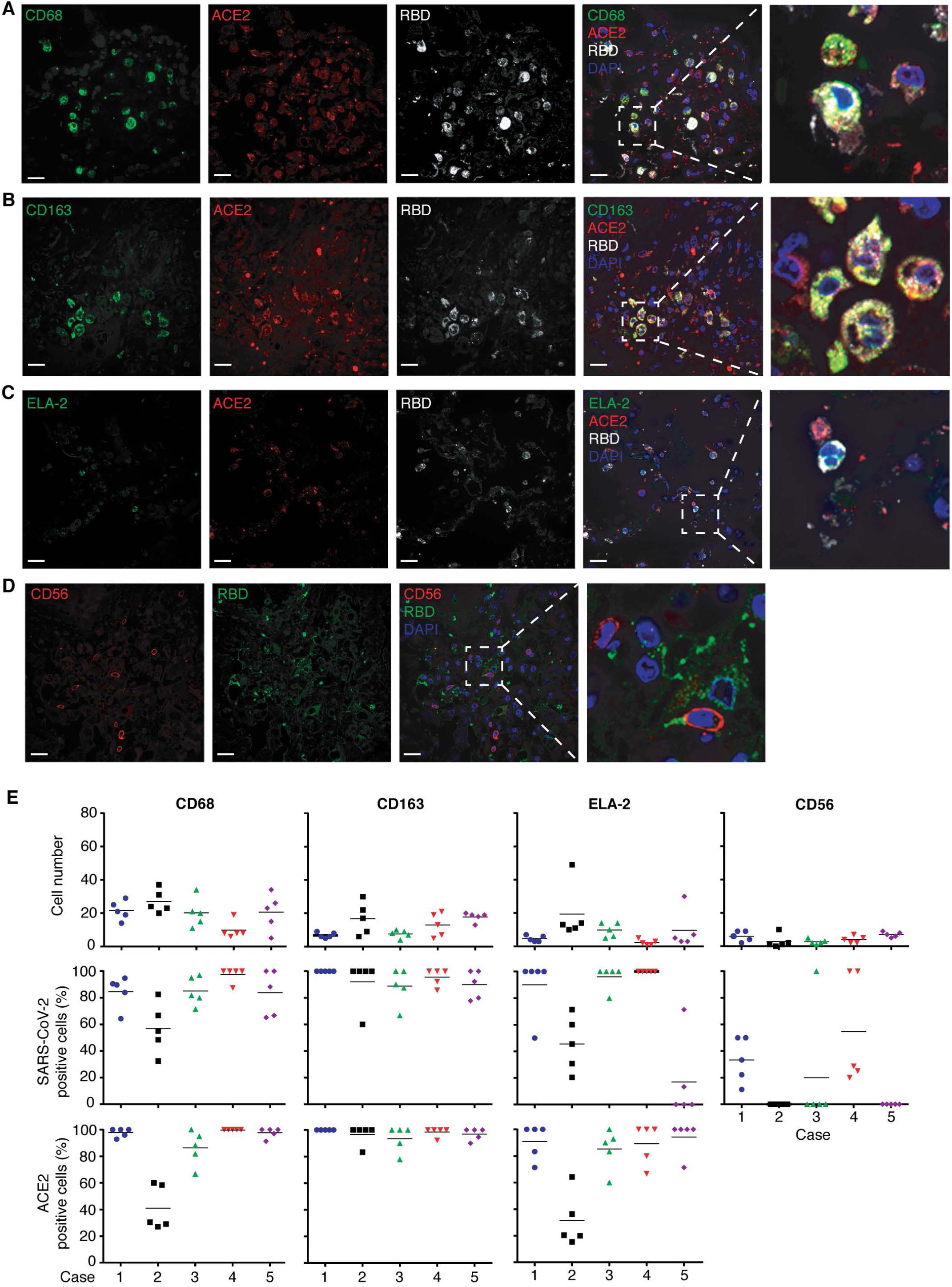
Representative images of multicolor immunofluorescence staining of ACE2, SARS-CoV-2 S1 protein (RBD) and markers of innate immune response cells in lung tissues from COVID-19 patients. Shown are CD68, a monocytic lineage marker (pseudo color green), ACE2 (pseudo color red) and S1 protein (pseudo color white) in case 4 (Panel A); CD163, a macrophage M2 marker (pseudo color green), ACE2 (pseudo color red) and S1 protein (pseudo color white) in case 5 (Panel B); Elastase 2 (ELA-2), a neutrophil marker (pseudo color green), ACE2 (pseudo color red) and S1 protein (pseudo color white) in case 4 (Panel C); CD56, a NK cell marker (pseudo color red) and S1 protein (pseudo color green) in case 5 (Panel D). Nuclei were stained with DAPI (pseudo color blue); and quantification of CD68^+^, CD163^+^, ELA-2^+^ and CD56^+^ cells in five different fields in each lung sample from all COVID-19 cases (figure S6). S1-positive and/or ACE2^+^ cells were counted in the same fields and shown as percentages of positive cells (Panel E).

Neutrophils, positive for elastase-2 (ELA-2^+^) protein, were extensively infected by SARS-CoV-2 (figure 3C and E, and S6C). The extent of neutrophil infection was positively correlated with ACE2 protein expression in all cases except for tissues from case 5, for which 96% of cells expressed ACE2, but only 19% had detectable SARS-CoV-2 infection (figure 3E). We detected SARS-CoV-2 infection in NK cells (CD56^+^) (figure 3D and S6D); however the percentages of infected cells were much smaller than other cell types examined, ranging from 0 to 40% (figure 3E).

Among the adaptive immune cells, B cells (CD20^+^) were found in low numbers in the lung specimens, but ACE2 protein expression and SARS-C0V-2 infection were positively correlated in these cells (figure 4A and F, and S7). Different types of T-cells expressing CD4 (figure 4B and F, and S8A), CD8 (figure 4C and F, and S8B) and CD3ε (figure 4D and F, and S8C) all co-stained with SARS-CoV-2 NC protein. In addition, HLA-DR, a marker of activated B and T cells, co-stained with SARS-CoV-2 NC protein (figure 4E and F, and S8D). Interestingly, CD4^+^, CD8^+^ or CD3ε^+^ T cells presented either as a membrane-associated pattern or as a dot-like organization pattern, possibly as the result of membrane rupture following SARS-CoV-2 infection (figure S8E).

**Figure 4:**
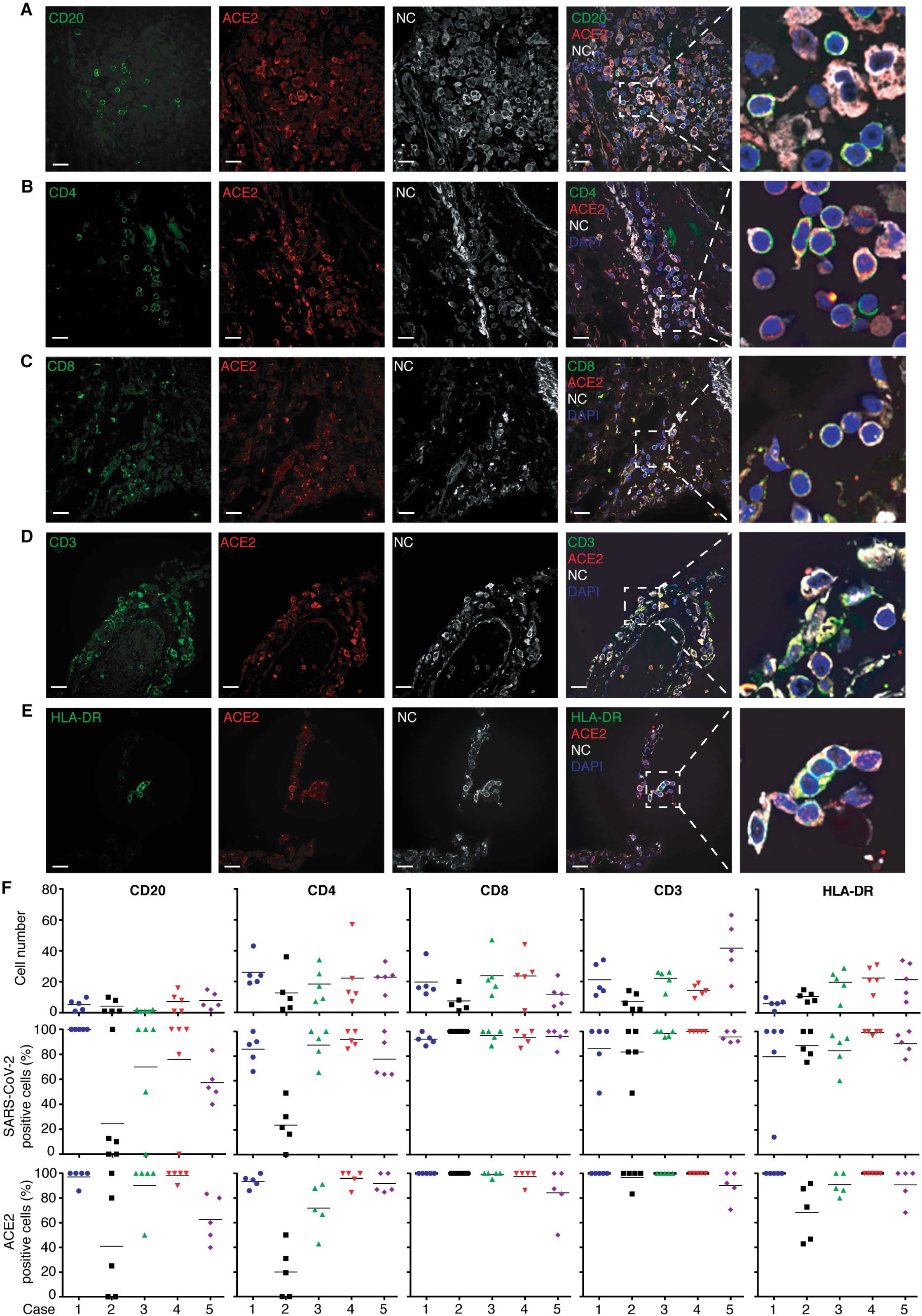
Representative images of multicolor immunofluorescence staining of ACE2, SARS-CoV-2 NC protein, and markers of B or T cells in lung tissues from COVID-19 patients. Shown are CD20, a B cell marker (pseudo color green), ACE2 (pseudo color red) and NC protein (pseudo color white) in case 5 (Panel A); CD4, a T helper cell marker (pseudo color green), ACE2 (pseudo color red) and NC protein (pseudo color white) in case 1 (Panel B); CD8, a cytotoxic T cell marker (pseudo color green), ACE2 (pseudo color red) and NC protein (pseudo color white) in case 1 (Panel C); CD3ε, a T cell receptor marker (pseudo color green), ACE2 (pseudo color red) and NC protein (pseudo color white) in case 1 (Panel D); HLA-DR, a B and T cell activation marker (pseudo color green), ACE2 (pseudo color red) and NC protein (pseudo color white) in case 2 (Panel E); and Quantification of CD20^+^, CD4^+^, CD8^+^, CD3ε^+^ and HLA-DR^+^ cells in five different fields in each lung sample from all COVID-19 cases (figure S7 and 8) (Panel F). NC-positive and/or ACE2^+^ cells were counted in the same fields and shown as percentages of positive cells.

IL6 is one of the most abundant cytokines detected in COVID-19 patients and its level is correlated with prognosis.^25-28^ In lung tissues from all 5 cases, we found expression of IL-6 in almost all cells examined, with or without SARS-CoV-2 infection (figure 5A-B and S9A-B). In general, the expression of IL6 was positively correlated with the number of infected cells (figure S9A).

**Figure 5:**
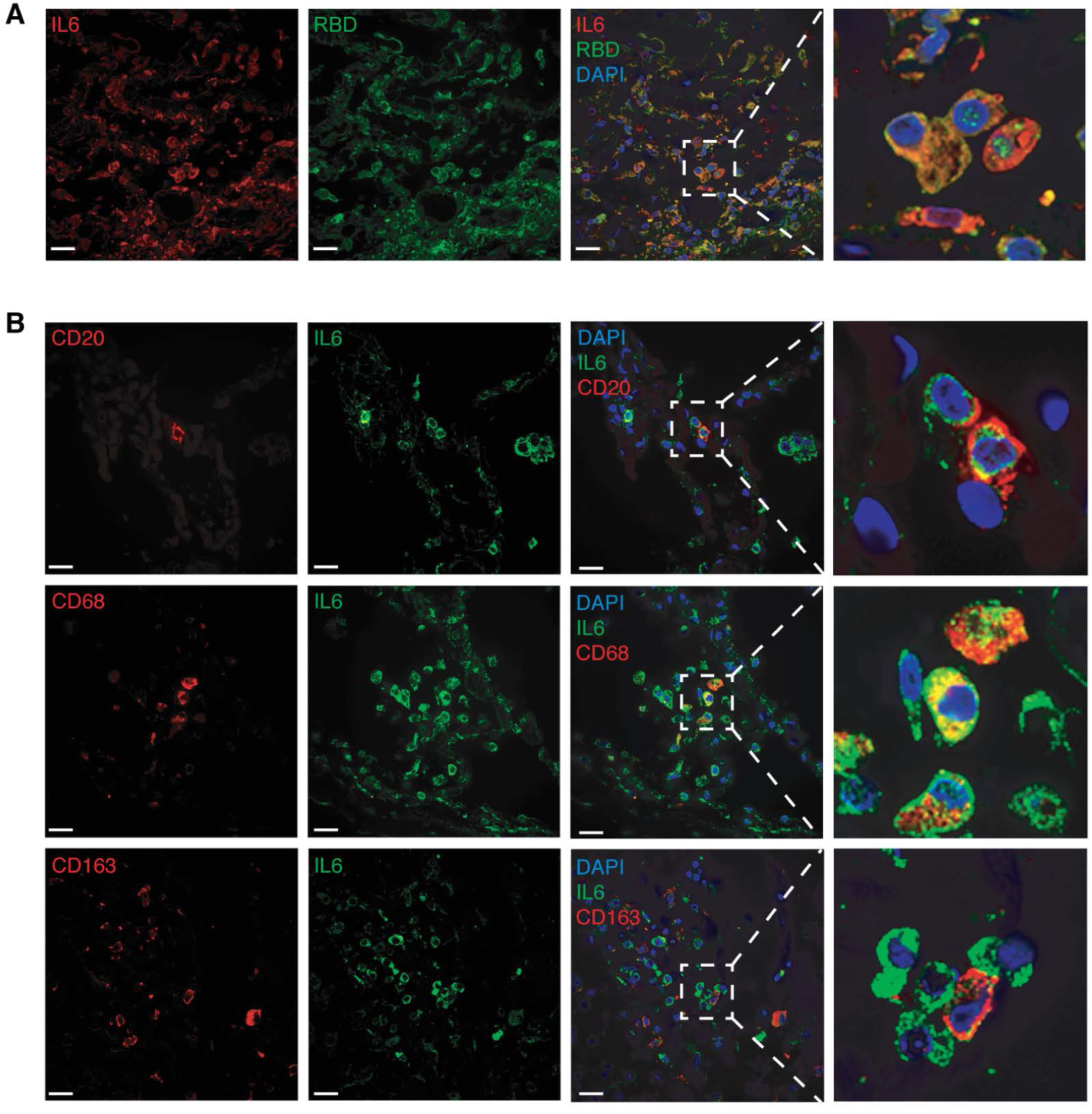
Representative images of multicolor immunofluorescence staining of IL-6, SARS-CoV-2 S1 protein (RBD), and cellular markers in lung tissues from COVID-19 patients. Shown are IL-6 (pseudo color red) and S1 protein (pseudo color green) in case 4 (Panel A); and IL-6 (pseudo color green), and cellular markers CD20 (pseudo color red), CD68 (pseudo color red) or CD163 (pseudo color red) in case 1 (Panel B). Nuclei were stained with DAPI (pseudo color blue).

## Discussion

Respiratory symptoms are a prominent complaint during most SARS-CoV-2 infections, and progressive respiratory dysfunction is a major feature of severe COVID-19.^1,2,6,38^ The results of our study present a direct visualization of the multiple cell types infected by SARS-CoV-2 from patients who died of COVID-19, and offer insight into the pathogenesis of the overwhelming damage found in lung tissues in fatal COVID-19 cases.

The expression of viral S1 or NC protein, as demonstrated by IHC and IF, indicated widespread SARS-CoV-2 infection in lung tissues, including multiple lung parenchymal cell types and multiple cell types involved in the immune response. These SARS-CoV-2 proteins were most abundant in specimens with the most histologic evidence of tissue damage. As expected, the extent of infection was positively correlated with the expression level of ACE2 protein. Notably, we also found SARS-CoV-2 infection in ACE2-negative cells, supporting a role for other possible receptors for viral entry into ACE-negative cells.

It is important to note that there may be multiple factors that might influence ACE2 expression during SARS-CoV-2 infection and should be considered as confounders. For example, ACE2 expression can be stimulated by IFNs.^17,18^ Furthermore, ACE2 may have a role in protection against severe acute lung failure, as has been reported in severe COVID-19 patients.^39^

We obtained direct evidence for widespread expression of ACE2 and extensive infection by SARS-CoV-2 among different cell types using multicolor IF staining for SARS-CoV-2 and ACE2 proteins in different pulmonary parenchymal and immune cells. Among lung parenchymal cells that were both ACE2-positive and SARS-CoV-2-infected included AT2 (HT2-280), ciliated (Tyr-α-tubulin), goblet (MUC5AC), club-like (MUC5B) and vascular endothelial cells (CD31^+^ or CD34^+^). Our findings are consistent with recent studies showing SARS-CoV-2 infection of ciliated, goblet and club cells by RNA-*in situ* hybridization (ISH);^40^ and infection of pneumocytes, ciliated, secretory and lymphomononuclear cells by IHC^23,41^ in lung tissues from COVID-19 patients. In *ex-vivo* culture, SARS-CoV-2 has been found to infect type I pneumocytes, ciliated, goblet and club cells as well as conjunctival mucosa.^22^

We detected ACE2 protein expression in different immune cells including CD68^+^ and CD163^+^ monocytes and macrophages, ELA-2^+^ neutrophils, CD56^+^ NK cells, and B- and T-cells; these findings are consistent with previous reports based on scRNA-seq studies.^7-10^ ACE2 expression detected by flow cytometry in T cells from lung tissues from COVID-19 patients has been reported by others.^42^ Notably, we found rates of SARS-CoV-2 infection approaching 100% for most types of immune cells, in contrast to a much lower infection rate in NK cells (figure 3E and 4F). Although previous studies have reported detection of SARS-CoV-2 proteins in macrophages by IHC,^20,43^ to our knowledge, our study is the first to demonstrate and quantify SARS-CoV-2 infection in different types of T cells and also in neutrophils. Our observation of SARS-CoV-2 infection of neutrophils was in contrast to the results of a recent study, which failed to detect any infected neutrophils.^42^

We simultaneously identified SARS-CoV-2-infected and ACE2-expressing cells in different parenchymal and immune cells in lung tissues from COVID-19 patients by multicolor staining. These observations are significant because infiltration and infection of immune cells such as macrophages are suggested as critical steps in the spread of SARS-CoV-2 infection to other organs^44,45^ and in the initiation of uncontrolled inflammatory responses.^46^ Furthermore, we have observed SARS-CoV-2 infection and damage of vascular endothelial cells together with the vast inflammatory infiltrations.

These evidences of endothelitis and direct viral injury suggest that endothelial cell dysfunction plays an important role in the genesis of thromboembolic events in SARS-CoV-2 infection.

SARS-CoV-2 infection has been proposed to cause compromised immune response by dysregulating the recruitment of immune cells.^47-49^ It has been reported that decreased levels of CD4^+^ and CD8^+^ T cells were associated with worsening COVID-19 outcomes,^48,50-52^ and there was evidence of activation of CD8^+^ T and NK cells as well as exhaustion of T cells in the lung tissues from COVID-19 patients,^53-56^ all of which could contribute to the increased proinflammatory or anti-inflammatory cytokines. In the lung tissues examined in this study, we noted a low level of CD20^+^ B-cells, and a lower level of CD8^+^ T as compared to CD4^+^ T cells. These results suggested a general immunosuppression in the lungs of COVID-19 patients. Most of the inflammatory infiltrates were characterized as CD68^+^, CD163^+^ and CD45^+^ cells. By contrast, we did not detect any FOXP3^+^ Treg cells, potentially supporting the T cell exhaustion theory,^54-56^ and the lack of Treg cells as a mechanism leading to failed control of inflammatory cells and the excess inflammation observed in COVID-19 patients.

The inflammatory cytokine IL-6 is highly expressed in COVID-19 patients, and elevated IL-6 levels have been associated with poor prognosis.^25-28^ However, the source of IL-6 in COVID-19 patients remains unclear. We detected a broad, increased IL-6 expression in all cell types and in lung specimens from all the cases we examined, and IL-6 expression could be correlated with the detection of SARS-CoV-2 proteins, as well as with the degree of tissue damage. Of note, our findings are consistent with previous studies reporting that patients with a high level of IL-6 and a poor prognosis also had decreased CD8^+^ T, NK and Treg cells.^53,54,57^

In summary, we performed a systematic analysis of SARS-CoV-2 infection in the postmortem lung tissues from patients with fatal COVID-19, providing an atlas of lung immunopathology of the disease. We found a broad tropism of SARS-CoV-2 infection in pulmonary parenchymal and immune cells. Finally, we observed evidence of activation of immune cells, exhaustion of B- and T-cells, and complete depletion of immune suppressive Treg cells, potentially contributing to the failure to modulate immune cell activation and response as well as inflammation. Further studies are required to delineate the mechanisms of SARS-CoV-2-triggered chemoattraction and immune exhaustion in the lungs of COVID-19 patients.

## Data Availability

N/A

## Contributors

SJG conceived and designed the study. SRDS, EGJ and WM designed and performed the experiments. AEPM, CB, ZG, MF, EMS and CCC collected clinical specimens and data. SD examined immunohistochemistry results. AG carried out part of immunohistochemistry. SRDS, EGJ, WM, HG and SJG contributed to data interpretation. SRDS and SJG wrote the first draft of the manuscript. All authors critically reviewed the manuscript, and approved the final manuscript for submission.

## Acknowledgments

We thank Drs. Yuan Chang and Patrick Moore for their insightful comments and suggestions, Elaine V. Byrnes and Paul Knizner in the Pitt Biospecimen Core for the technical support. This work used the UPMC Hillman Cancer Center and Tissue and Research Pathology/Pitt Biospecimen Core shared resource, which is supported in part by award P30CA047904. This study was supported by UPMC Hillman Cancer Center Startup Fund and Pittsburgh Foundation Endowed Chair in Drug Development for Immunotherapy to S.-J. Gao.

